# A Note on the Effects of Digital Primary Health Care on Utilization: Concepts, Evidence, and Descriptive Analysis of Non-Experimental Register Data from Sweden

**DOI:** 10.1101/2022.06.28.22277001

**Authors:** Björn Ekman, Hans Thulesius, Jens Wilkens, Eva Arvidsson

## Abstract

Digital technologies for health care may lower costs while enhancing access to services. However, concerns have been raised that digital care may lead to over-use of services and not be as effective as in-person visits. Previous studies have found varying effects across different contexts, study designs, and outcome measures. This study contributes to the emerging evidence on the effects of digital care on primary care utilization by developing a conceptual model for primary care use and then comparing the effects of digital primary care with in-person visits and telephone contacts. Register data from Sweden over a two-year period (2017-2018) in a sample of patients diagnosed with an infection are used to describe the effects. Findings show that the majority of patients require a single consultation across either model of care. A relatively small share of patients makes multiple consultations per episode of care both across and within models of care. Compared with in-person visits and telephone contacts, digital care is associated with fewer consultations per episode of care and involve lower rates of laboratory tests and antibiotic prescriptions. Digital care is provided by a physician to a larger extent compared with the other models of care in the current sample. Further analysis will be conducted to establish any causal effects of digital primary contacts on identified outcomes.

## Introduction

Over the past decade or so, digital technologies for providing health care over the Internet have expanded rapidly across many countries and regions (OECD 2017, Hamilton 2019). While specific technologies vary, they involve some form of remote patient consultation that represent both an advancement of existing technologies, such as the telephone (and letter), and a replacement of traditional models of health care in the form of in-person, office-based visits. In the current literature, these models are variously called, among other things, direct-to-consumer (DtC; largely in the U.S. context), e-visits, virtual visits, and telemedicine (Jayabalasingham 2020). While their names and specific designs differ across systems digital technologies have enabled new forms of providing health care that bridge the gap between the patient and the provider both in terms of time and space. This is particularly relevant in primary health care (also referred to as general practice (GP) and family medicine; ambulatory care for both new non-emergency illnesses and some chronic conditions) where digital models of care have enhanced access to services for many patients (Baird and Nowak 2014, Nilsson, Sverker et al. 2021).

The possibilities that digital health technologies give rise to have received significant policy attention across most health systems. The interest in being able to provide primary care remotely by means of digital technologies has been particularly strong during the Covid-19 pandemic when in-person visits have been restricted (Fahy and Williams 2021). In parallel, questions have been raised as to the risks that the expansion of various forms of digital health care can bring, including that of excessive utilization of services beyond what is medically motivated and of over-use of antibiotic pharmaceuticals (Iacobucci 2018). Suggestions have been made that digital care does not or only partially substitute for in-person care but rather leads to an increase in the overall utilization of services (Cheung, Leung et al. 2019). Concerns have also been raised that digital care is not as effective as in-person care for certain conditions that require the ability to physically examine the patient. Indeed, the World Health Organization has issued a special statement and guidelines on the appropriateness of digital health care (WHA 2018, WHO 2019).

Sweden represents an example where the increase in the provision of digital primary health care has been particularly strong over the past five to six years. Since 2016, a group of private, for-profit digital-only providers of primary care services have gradually expanded their operations in a predominantly public system (Ekman, Thulesius et al. 2019). In Sweden, health care is organized, financed, and largely provided by the 21 health care regions. With respect to primary care, the regions either operate their own clinics or contract with private providers of such services. Up until the mid-2010s, the vast majority of primary care consultations were made by in-person visits or, to a lesser extent, by telephone and letter (mostly for follow-up consultations and advice). While digital care still constitutes a minority of total primary care consultations, their overall share is today most likely one of the largest in the OECD group of countries (Hashiguchi 2020). The development of digital care in Sweden and its broader impacts may therefore be of interest to other countries.

However, the expansion of digital primary care has also in this case led to a discussion of the appropriateness of these kinds of services. The main argument for their existence is that they increase access to care. In the current case, this is broadly seen as positive as Sweden has struggled with compromised accessibility to primary care for a long time (Anell, Glenngård et al. 2012). The main counter argument is that these services lead to over-use of care and thereby distort the allocation of resources. Regardless of the strength of these arguments, the introduction of new models of care and their scaling-up need to be motivated by convincing evidence that they represent value for money.

Investigating the extent to which digital care affects overall utilization requires an understanding of the mechanisms whereby the effects would operate. It is likely that these are found both on the demand and on the supply side of health care utilization. On the demand side, patients may perceive digital care to be at least as effective as traditional, in-person care. Patients would then choose the least costly type of care in terms of financial and time costs. For example, digital care may be seen as a more convenient model of care.^1^ On the supply side, digital care may be less effective than in-person services and require more intense levels of care in the form of more consultations per episode of care. Alternatively, it may require a different provider mix and a higher use of laboratory testing compared with in-person care. Yet another possibility is that if symptoms remain after the use of digital care, patients may switch to in-person care. In all cases, more resources would be required on a per case basis. Understanding the extent to which there is a difference in the effectiveness of alternative care models is therefore important for policy development and decisions on resource allocation.

However, comparing the effects of different models of care is difficult for two main reasons. First, in the absence of an experimental study design, in which patients are randomly assigned to receive care in either of the alternative models, observed differences in outcome could be the result of selection bias as patients self-select to the particular model of care. And second, identifying and estimating the particular outcome measures of the relevant effects of health care, including primary health care, is challenging (Soumerai, Starr et al. 2015). The difficulties are largely due to the complexity of health care and the lack of information on relevant aspects of the health care process, including final treatment outcomes, such as QALYs^2^ (Black 1996).

Against this general background, the overall purpose of this study is to contribute to the current evidence on the effects of digital care on the utilization of primary health care services. Using consultation level data of three primary care models, in-person visits, digital contacts, and telephone/letter contacts, the empirical analysis aims at estimating the extent to which there is a difference in the effects of the alternative models of primary care on a set of measures across three dimensions of primary care: the intensity (or quantity) of care per episode; the possibility of a switch of care model; and the content of care (see the Methods sub-section for specific outcome measures).

A general finding of the analysis is that the majority of patients make a single consultation per episode in either model of care. A share of the patients makes multiple consultations per episode, the majority of whom use the same model of care. Switches between models of care occur across all models of care, albeit to a very limited extent. The digital model of care differs most strongly with respect to the content of care as almost all contacts are made with a physician, fewer laboratory tests are conducted, and less antibiotics are prescribed compared with the other models of care.

The next section presents an analytical framework to guide the analysis of the study and to support the interpretation of the findings. The subsequent section provides an overview of the existing evidence on the effects of digital care on the utilization of primary care. This is followed by a description of the data and the methods used in the study. The findings of the analyses are presented in the subsequent section, and these are discussed in the concluding section. Additional tables and graphs are provided in the Annex. The study has been conducted as part of a research project funded by the Swedish Research Council for Health, Working Life, and Welfare (Forte; grant number 2018-00093). The research project received ethical approval from the Swedish Ethical Review Authority (2019-01500; 2019-03-20).

### Evaluating the effects of digital primary care: an analytical framework

The introduction of new forms of health care is likely to affect existing models of care, which in turn will affect the overall health system and its performance, both in terms of efficiency and distribution. However, new models of care are rarely implemented in a controlled randomized manner. This makes it difficult to obtain a control group for the causal evaluation of the effects of the new model. Consequently, empirical analyses of the effects of alternative models of care need to employ non-experimental methods. Regardless of the particular empirical method understanding these effects on selected outcomes requires the existence of a theoretically sound framework to support the analytical approach. Importantly, a theory-based analytical framework may assist in capturing the data generating process in empirical analysis and in interpreting the findings of the analysis.

While alternative such frameworks have been suggested across various disciplines, such as sociology, political science, and public health, the (neoclassical) economics view is that observed market outcomes are the result of the interaction between demand and supply factors. On the demand side, the individual is faced with a choice-set, the outcome of which depends on her preferences, expectations, and budget constraints (Mwabu 2017, Phelps 2018, McPake, Normand et al. 2020). Technically, the individual choses a consumption bundle to maximize her utility given the budget constraint.^3^ On the supply-side, alternative models of care may contain different levels of human and capital factors to produce a unit of care. It is likely that the production function differs across different models of care.

In primary care, unlike some other forms of health care, including emergency care and specialized in-patient and ambulatory care, it is the decision of the individual to initiate the care process. As such, primary care is a demand driven choice process. More precisely, when experiencing certain symptoms, the individual is faced with a choice between two types of care, namely provider-directed care (whereby the provider acts as the individual’s agent in terms of scope and content of the care) or self-care (in which the individual acts as her self-agent and which may contain certain technologies and activities, such as non-prescription drugs and rest). The first type of care is generally more costly in terms of direct and indirect costs compared with the second type of care. On the other hand, provider-directed care may be viewed as more effective compared with self-care. These differences are likely to affect the individual’s choice with respect to which type of care to seek.

Figure 1 (Panel A) describes the framework for analyzing the primary care process by illustrating the individual’s choice options at the various levels of this process.

**Figure 1.**
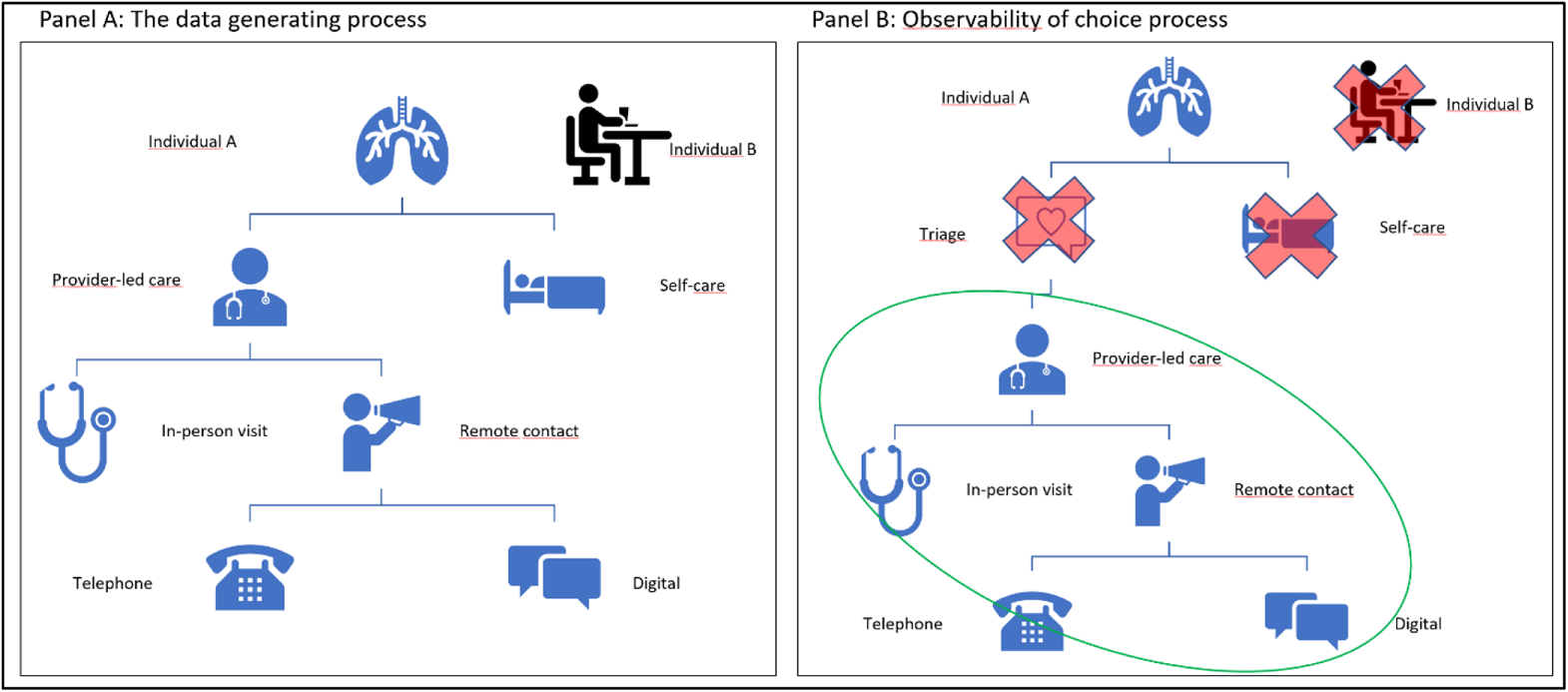
The primary care choice process. Source: Authors.

As symptoms appear, Individual A makes a choice between the two main models of care, provider-led care and self-care. The former type of care comes in two forms, in-person visit or remote contact, both of which are externally taken as given by the individual.^4^ In turn, remote consultations are of two types, digital or telephone/letter contacts. Importantly, it is the individual’s choice which type of care to seek, i.e. the individual self-selects into the various states in Figure 1 Panel A. Indeed, it is this self-selection process that complicates the causal analysis of the effects of alternative models of care on selected outcomes.

The ability to make causal inferences depends largely on the observability of the choice process illustrated in Figure 1, Panel B. First, at any given point in time, the individual can only be observed in one state of being. This is the core issue in any impact evaluation effort. Second, data are rarely available to distinguish between those who experience symptoms during a particular period (Individual A) and those who do not (Individual B). The process that drives the realization of these two states of being in a population is largely unknown but can be assumed to contain substantial randomness and idiosyncrasies. Depending on the purpose of the empirical analysis, the lack of data on this part of the choice process may affect the ability to make inferential conclusions. Furthermore, if there are factors that drive this process that also affect the choices made with respect to health care, this lack may be relevant for the interpretation of empirical results.

Another issue that is also shown in the figure may be more serious as it directly affects the analysis. Information about the individual’s choice between physician-led care and self-care may also not be readily available. Indeed, most empirical studies (including this one) on the effects of digital care have used register data on those who actually made the choice to seek physician-led care. The unobservability of this part of the choice process may affect the ability to draw conclusions about the causal effects of digital care on the overall utilization of services, including the extent to which digital care substitutes for other types of care, of which self-care is one. The figure highlights the part of the primary care choice process on which this study has data (encircled in green in Panel B).

The analysis of health care utilization is complicated by factors on the supply side. First, in both in-person and in digital care most patients are screened in a triage process before being seen by a physician. In the in-person case, this process involves the need to speak to a nurse over the phone who then decides whether further physician-led care is necessary or not. Those consultations that do not result in an appointment or advise to return if symptoms persist are commonly not registered in the data. In the case of digital care, the triage system is automated and information introduced by the patient is analyzed by artificial intelligence-driven algorithms that inform clinical decisions.

Once a consultation has been initiated, it is, implicitly, a joint decision by the individual and the provider whether to pursue further care or not. The informational advantage of the provider may affect the individual’s decision to seek further care of some type, both in terms of scope and content. This is referred to as supplier-induced demand (SID) in the health economics literature and has been found to be real in primary and other types of care (Longden, Hall et al. 2018). In empirical analysis of primary care use, the behavior of providers is largely unobserved *ex ante*. Instead, the behavior of providers is derived from the *ex post* observations of the care process, including on diagnosis, examinations, prescriptions, and (occasionally) referrals.

As noted, physician-led care can be assumed to be more costly compared with self-care. However, the introduction of digital primary care has led to, among other things, a reduction in the transaction costs associated with utilizing services. In particular, transportation and time costs on the part of the patient are generally lower in digital care (Dullet, Geragthy et al. 2017, Ekman 2017). The reduced access costs can be assumed to affect the individual’s preferences over both physician-led care and self-care and over in-person visits or remote contacts as it affects the relative prices of care.

However, two issues may arise from this situation. First, if the digitally based, physician-led care mainly substitutes for self-care and not for in-person, physician-led care, the level of physician-led care that may not be medically motivated would increase. The introduction of digital care would then lead to a situation where more unnecessary care is provided. While measuring unnecessary care is possible, it requires detailed patient record data of sufficient magnitude (Liu and Mills 1999).^5^

The second issue of relevance is if digital care substitutes for office-based care. This may occur for two reasons. First, the individual’s preferences may be affected such that utilizing one form of care at time *t* may affect her choice at time *t + 1*, regardless of when the subsequent consultation takes place. And second, digital care may be less effective compared with in-person care and, all else equal (i.e. for the same diagnosis and time period), may require more consultations per episode of care or a different technological input mix in terms of human and capital (e.g. physician-led care rather than nurse-led care, and laboratory investigations).

The framework outlined above identifies a number of ways that the effectiveness of digital care may be different compared with traditional, in-person care, including in the overall use of care per episode, the probability of changing from one model of care to another in subsequent consultations, and in the content of the care. Some of these mechanisms have been investigated in previous analyses as described in the next section.

### Overview of current evidence

The evidence on digital technologies for health care has increased rapidly over the past decade or so. It also involves a wide range of technologies across various types of health care. This section provides a broad overview of the recent evidence on the use of digital technologies for providing primary care. It includes a selection of relevant systematic reviews of the evidence and of specific studies of particular relevance. It concludes with a summary assessment of the existing evidence to describe the contribution of the current study.

### Systematic reviews

As the applications of digital care models have proliferated, so has the number of studies on their effects. Importantly, studies on the impact of digital care on, among other outcomes, levels of utilization have been the subject of some systematic reviews (including of systematic reviews) and meta-analysis. In 2006, Chaudry and colleagues systematically reviewed the evidence on, among other things, the extent to which “health information technologies” (HITs) had an effect on health care utilization Chaudhry, Wang et al. (2006). They found that out of 11 studies included in the review, eight reported decreased utilization as a result of the use of a technology, including for decision-support at the clinical level. Utilization rates decreased by between 8.5 and 24 percentage points. The types of services included were laboratory testing and radiology and mostly concerned hospital types of settings.

In a systematic review and meta-analysis of systematic reviews, Elbert and colleagues (2014) found that “eHealth interventions” are generally effective or cost-effective and that overall “the evidence is promising” (Abstract). However, the evidence with respect to family medicine was found to be smaller compared with other specialties. Finally, Zanaboni and colleagues (2018) systematically reviewed systematic reviews of the evidence on “internet-based health interventions” and found, among other things, that the evidence on the effects on utilization was mixed with some studies reporting decreased utilization while others had found the opposite effect. They also concluded that the quality of the evidence was low.

### Discrete studies

With respect to specific studies on the effects of digital care models on utilization rates of primary care, a handful of studies have found varying impacts. An early contribution to this issue is Angstman and colleagues (2009). They evaluated the impact of virtual visits (termed “e-Consults”) on the frequency of return visits for a small sample (n = 728) of primary care patients in the U.S. They found that the share of return visits for any reason within 2 weeks of the consultation was 38 percent for users of digital care compared with 28 percent for patients who had made an in-person visit. After adjusting for a set of observable factors, the odds of an early return visit for any reason after a digital visit were 1.88. They concluded that digital care may lead to more follow-up visits.

Mehrotra and colleagues (2013) analyzed claims data collected from four primary care clinics in the U.S. to assess differences between e-visits and in-office visits for respiratory tract infections (RTI) and urinary tract infections (UTI). They found no difference in the rate of follow-up visits within three weeks between the two models of care for either of the condition. They did find differences with respect to ordering tests and prescribing antibiotics. In particular, there was a higher test rate for UTI for in-office visits compared with e-visits, but no difference for RTI. There was a significantly higher rate of antibiotic prescriptions for e-visits, although in both models of care the prescriptions were seen as guidelines concordant. In a similar study, Uscher-Pines and colleagues (2014) analyzed claims data from a state level health plan in the U.S. (n = 75,000 users) to describe users of care and the effects of telemedicine on overall utilization of services. In addition to describing the sociodemographic characteristics of the sample, they found that telemedicine patients had lower utilization rates compared with users of in-person care. Likewise, North and colleagues (2014), in a study of 2,357 individuals in the U.S., did not find any significant differences of in-person primary care visits before the introduction of an “e-consult portal” compared with after the introduction. Also in a U.S. context, Gordon and colleagues (2017) report similar results in a study of virtual primary care contacts where they compared follow-up rates and laboratory testing and image rates to those of in-person visits for a defined episode of care of three weeks after the index visit. They found that virtual visits had similar follow-up rates and lower prescription and image rates compared with in-person visits.

In 2017, Ashwood and colleagues (2017) used a large sample (n ∼ 300,000 of which there were n = 981 telehealth visits matched to n = 1,962 in-person visits) to study the questions of whether digital primary care leads to less per episode spending and to fewer in-person visits, in the U.S. They used statistical modeling involving difference-in-differences (DiD) and propensity score matching (PSM) to learn that the per episode cost of care was smaller for digital contacts compared with in-person visits and ER-visits, but also that overall utilization of services increased as a result of digital care. Specifically, digital contacts replaced around 12 percent of in-person visits with the balance representing new utilization that would not have taken place in the absence of digital care (a substitution rate of 12 percent).

Bavafa and colleagues (2018) report partly similar findings in a study using difference-in-differences (DiD) regression estimation on a sample of around 100,000 patients using e-visits in a U.S. context between 2008 and 2013. They found that such visits lead to increased use of in-person care and that this came at the expense of new users of services. Johnson and colleagues (2019) estimated differences between virtual visits and office visits for patients diagnosed with sinusitis within a primary care system in the U.S. They found, among other things, that virtual patients were more likely to make a follow-up visit within 24 hours and within 30 days of the initial visit.

Ray and colleagues (2019) investigated trends in acute direct-to-consumer care of children using data from a national health plan in the U.S. They found, among other things, that compared with children who had visited an in-person clinic for acute care, children who had used DtC care had higher rates of urgent care and emergency visits during the study period of 2011-2016. A recent study by Li and colleagues (2021) took a slightly different approach by looking specifically at the rate of follow-up consultations after an initial direct-to-consumer contact or an in-person visit in a primary care system in the U.S focusing on respiratory infections. In particular, they identified utilization of care during a 7-day episode of care period. The authors found that an initial digital contact led to a follow-up consultation of some type (digital, in-person, urgent care clinic, or emergency department) in 10.3 percent of cases compared with 5.9 percent for in-person visits. Using statistical chi-squared tests, this difference was found to be statistically significant.

Studies on the effect of digital care on utilization have also been conducted on samples of patients diagnosed with a chronic condition. Liss and colleagues (2014) used interrupted time-series analysis to assess the effects of a secure messaging and telephone service on in-person visits among a sample of persons diagnosed with diabetes in a nursing home setting in the U.S. They found an increase in office visits after the implementation of the service. Berkhof and colleagues (2015) randomly assigned patients diagnosed with COPD to telemedicine care and a control group receiving usual in-person care (n=101). They found that patients in the control group made fewer visits to see a pulmonologist during the study period. Valdivieso and colleagues (2018) used a matching approach to a small number of elderly patients with multiple chronic diagnoses in Spain. They report a small positive effect of telehealth use on in-person primary care visits. Using partly similar types of data and analytical approach (quasi-experimental methods with n = 35,854; n = 519 matched individuals), Sha and colleagues (2018) addressed the question of whether digital care lead to a reduction in the use of in-person specialized (neurology) services in the U.S. The authors found that virtual contacts were associated with a reduction in in-person visits of 33 percent but also with an 88 percent increase in combined virtual and in-person visits. They also report that to offset one in-person visit required 3.5 virtual contacts in the sample of neurology patients. Based on their findings, they find a substitution rate of 29 percent.

### Evidence from Sweden

In the Swedish context, two recent studies using similar data as the current study, have examined the effects of digital primary care on overall utilization of services. Using register data from 2016-2018 from one Swedish region, Ellegård and Kjellsson (2019) modeled the effect of digital primary care on aggregate primary care visits and emergency visits. They found that one digital care contact led to 1.46 more primary care contacts with a physician and to 0.46 more in-person visits. They concluded that digital care does not “alleviate” the regional primary care system.

Using register data from two other Swedish regions combined with background data on patient characteristics, Ellegård and colleagues (2021) studied the effects of direct-to-consumer care on overall primary care utilization. By including observations from before the introduction of digital care and focusing on patients 18 years of age affected by a change in user-fees (as the identification method), they adopt a fuzzy difference-in-discontinuities (DiDi) regression design to control for unobserved confounding factors. They interpreted their findings such that digital care had a causal impact on in-person primary care visits. Specifically, the authors make the counterfactual claim that one digital contact resulted in 0.5 in-person visits “that would not have taken place if DtC was not available” (p. 3; a substitution rate of 50 percent).

Finally, in a recent study, Entezarjou et al (2022) used data from a set of primary care clinics in southern Sweden to investigate the extent to which users of a blended digital-physical care model (asynchronous eVisits; n = 1,188) made more follow-up visits compared with an initial in-person visit (n = 599) during a two-week follow-up period. They find that patients using the eVisit model of care had a higher proportion of a physical revisit within 48 hours, but not over the entire follow-up period.

### Assessment of current evidence

As new and innovative approaches to delivering services to patients are rarely implemented in a controlled or randomized manner, researchers have applied various types of statistical analyses, including quasi-experimental methods to evaluate the effect of alternative models of care. These methods cover both case-control and before-after designs. Where possible, these designs have been strengthened by using matched samples and difference-in-differences (DiD) or some type of discontinuity design (or a combination thereof). The findings reported from these studies are mixed, with effects seemingly going in different directions or no effect at all. Importantly, no study has been identified that explicitly addresses the question of whether digital care substitutes for self-care. Rather, as indicated above, results have been interpreted to suggest that this may be the case by, for example, Ashwood et al. (2017) and Ellegård et al. (2021) despite the unavailability of data on this part of the choice process.

Moreover, the studies of the impact of digital care on utilization can be broadly divided into two groups. One group of studies (Ashwood et al. (2017), Sha et al. (2018), Ray et al. (2019), and Ellegård et al. (2018, 2021)) have used frequency of consultations during some unspecified period after the introduction of the digital care model as their outcome measure. The other group of studies have focused on the effects within a specified episode of care of some length (between seven and 30 days) (Angstman et al. (2009), Ashwood et al (2017; with respect to costs), Gordon et al. (2017), and Li et al. (2021); Shi et al. (2018) focused on antibiotic prescription and identified episodes of care).

While both approaches to assessing the impact of digital care have their place in the evidence base, they would appear to capture different data generating processes. Measuring the universe of consultations after the introduction of the new model of care and then accruing any change in this outcome measure to the (utilization of the) digital model provides a broad, general measure of the (potentially causal) impact of the utilization of such care. It is not clear, however, if the change is due to the effectiveness (or lack thereof) of digital care or to latent preferences on the part of some share of the users. For example, a person may have made a digital consultation in, say, February, and then an in-person visit in September of the same or even following year. It is unlikely that the second consultation was medically motivated by the outcome of the first consultation for these types of conditions. Such studies would consequently fail to capture any difference in the effectiveness of the alternative models of care. The first consultation may, however, have affected the person’s preferences with regard to type of care in subsequent health care decisions. As discussed above, whether this is in fact the true data generating process is not known due to the unobservability when using register data.

This approach also puts strong demands on the available data. First, the data need to be able to control for any secular trend in utilization. Indeed, in most cases, such a trend is present, including in Sweden where observed utilization of primary care has been going down over the past decade or so. Second, the data generating process cannot be affected by an administrative change in the registration of consultations in the database. Indeed, the very introduction of a new model of care may have led to such a change. And third, the data truly need to capture the “universe” of consultations, including care provided outside of the regular primary care system, such as that covered by private health care insurance or by employment- or school-based health care services. Moreover, while it is true that the causal path would appear to go from digital care to in-person visits, the analysis of a reverse relationship would seem to be in place.

On the other hand, analyzing the impact of (digital) care within a certain episode of care of some length (for the same condition) would appear to provide a more direct estimate of the effectiveness of the particular care model. For example, if it can be shown that, on average, digital care requires a larger number of consultations per episode compared with in-person visits, the implication would be that digital care is not as effective as the alternative. Whether this also translates into higher spending depends on the unit cost of each type of model of care (Ashwood et al., 2017). Similarly, if digital care requires a larger number of laboratory tests or other investigations, or if a larger share of cases is handled by physicians, compared with in-person care, the implications would be that the potentially positive net effect of digital care would be smaller than first assumed. Generally, any approach that fails to account for the data generating process would be unable to provide clear and relevant policy guidance. As noted above, analyzing the use of services per episode would appear to more accurately capture the mechanisms generating the relevant values of the outcome measures. The current study adopts the “episode of care” approach to investigating the effectiveness of digital care on primary care utilization using a purposive dataset from Sweden.^6^

## Data and Methods

### Data

The data were collected in 2019 and cover a 730-day period from 1 January, 2017 to 31 December, 2018 (resulting in both right- and left-censored data). The data sources include Statistics Sweden (demographic and socioeconomic indicators of individuals), the National Board of Health and Welfare (in-patient care and pharmaceutical prescriptions), and seven regions (out of 21 regions; Jämtland Härjedalen, Örebro, Stockholm, Östergötland, Halland, Jönköping, and Kronoberg; primary care databases with utilization data over the sampling period). To reduce the problem of variations in the case-mix across care models the study focused on collecting data on primary care consultations for patients that had been diagnosed with an infection diagnosis, including respiratory, urinary, and skin- and soft-tissue infections. The study adopts the identification of these diagnoses suggested by Cronberg et al (2020). These conditions are common in both digital and in in-person primary care in most geographic contexts generating substantial overlap across models of care.

In the current study, the unit of analysis is the model of care that provide three types of consultations: in-person visits; digital contacts; and telephone or letter contacts. Figures A. 1 and A.2 in the Annex show the development of consultations over the sampling period; understanding the possible association between the observed changes is the aim of the study. Table 1 presents the distribution of consultations across the three diagnosis groups by type of consultation. Figures A.3 and A.4 in the Annex show the overall distribution of the sample of consultations.

**Figure 2.**
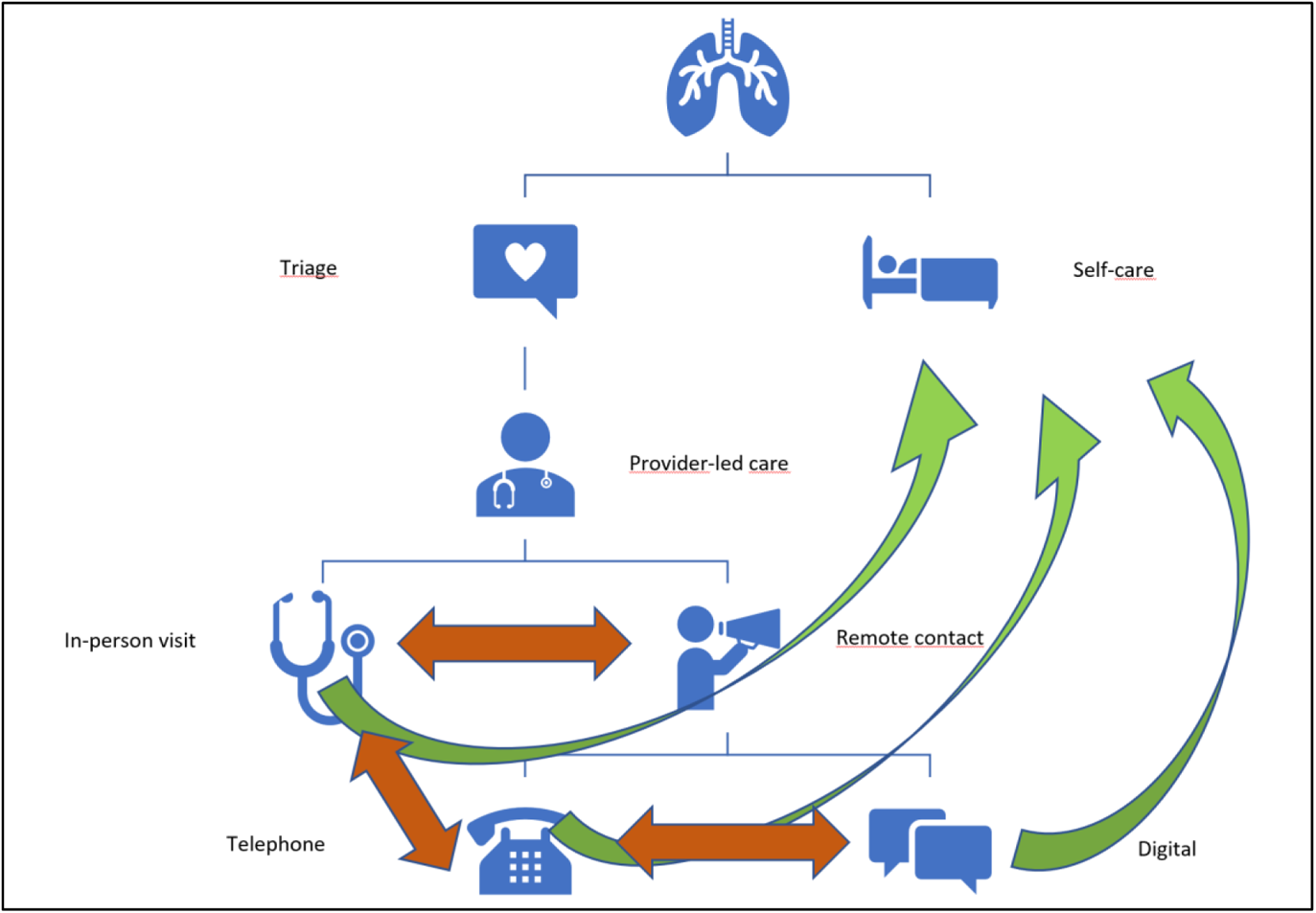
Models of primary care and their relationship in utilization.

**Table 1.** Descriptive statistics of study sample; frequency and percent

|  | Visit | Type of consultation |  | Total |
| --- | --- | --- | --- | --- |
|  |  | Digital | Telephone |  |
| Type of infection |  |  |  |  |
| Respiratory tract infection | 904,188<br>63.96% | 76,595<br>52.26% | 55,285<br>66.77% | 1,036,068<br>63.05% |
| Urinary tract infection | 160,971<br>11.39% | 32,115<br>21.91% | 18,313<br>22.12% | 211,399<br>12.87% |
| Skin- and Soft-tissue infection | 348,616<br>24.66% | 37,842<br>25.82% | 9,207<br>11.12% | 395,665<br>24.08% |
| Total | 1,413,775 | 146,552 | 82,805 | 1,643,132 |

Source: Digital primary care study, DPC; consultation level data for sampling period. Telephone includes letter contacts.

In the current sample, in-person visits make up around 86 percent of the total number of consultations. Digital contacts constitute around eight percent and telephone or letter contacts around six percent of the total. In-person visits include all face-to-face consultations between the patient and the provider (e.g. physician, nurse, or other health care professional). Digital contacts include consultations between the patient and the provider by means of some internet-based digital technology, such as encrypted video or text communication using a computer, tablet, or mobile phone. During the sampling period, the majority of those consultations were provided by a group of private, digital-only providers operating through existing primary care clinics in the Region of Jönköpings län (Ekman, Thulesius et al. 2019). These consultations were made by patients living in any of the Swedish regions.^7^ Additional digital contacts were provided by primary care clinics in the selected regions. Telephone and letter contacts are registered as one type of consultation in the regional databases and are made exclusively with the same clinics to which the in-person visits are made.

Table A. 1 in the Annex describes the sample at the individual level. After removing observations without complete values for the main variables of interest (including contact date and diagnosis) the sample consists of 673,721 individuals aged 15 to 110 years old, around 59 percent of whom are women. In the current sample, women are generally slightly older compared with men, report a lower annual income, and a lower value on the Charlson comorbidity index (CCI), a measure of overall health (Charlson, Charlson et al. 2008, Ludvigsson, Appelros et al. 2021). Relatively more women have a university education and are unmarried, while relatively fewer women have been diagnosed with a chronic illness, such as diabetes, COPD, or hypertension. There were small differences between men and women in the sample with regard to country of birth and employment status.

In addition to these individual level indicators, the data also contain a set of context indicators, including a care-need index (CNI; mean = 1.05, s.d. = 0.14, min = 0.63, max = 1.58) at the municipality level (Sundquist, Malmström et al. 2003), the share of fee-for-service (FFS) in total provider reimbursement at the regional level (mean = 16 percent; Lindgren (2019)), and the primary care user-fees in effect during the study period, also at the regional level (median 200 SEK; range: 0 – 300 SEK; 100 SEK = approx. US$9.5, June 2022; (SKR 2020)).^8^

### Methods

To investigate the effects of digital care the study employs statistical analysis to describe utilization along the three defined dimensions of care, namely intensity of care, change of care model, and content of care. Specifically, the study estimates the following outcome measures by model of care:

A. Intensity of care:

1. The number of consultations of the same type of care per episode of care
2. The share of consultations that involve a single consultation per episode of care
B. Change in model of care

1. The number of follow-up consultations that involve a switch from one type of model to another; both within an episode of care and across the study period
2. The share of follow-up consultations that involve a switch from one type of model to another; both within an episode of care and across the study period
C. Content of care

1. The share of consultations that are provided by a physician
2. The share of consultations that involve a laboratory investigation
3. The share of consultations that include an antibiotic prescription

These measures are presented in tables by dimension using unweighted frequencies and shares (percent) of total number of consultations by model of care. The tables show the main results using the total sample. In subsequent analyses, the measures are estimated for separate groups, including across the three types of infection diagnoses. With respect to the issue of the intensity of care, these measures may be sensitive to an excessive number of consultations by a relatively few number of patients, in particular if made by the same type of care. In the sensitivity analysis, the sample is adjusted to mitigate any such effect by removing individuals with a very large number of consultations.

In addition, there may be differences across the models of care as a result of differences in case-mix between the care models. To obtain a better understanding of the potential impact of such factors, differences between the types of consultations across patient characteristics are analyzed by means of statistical tests. Differences across continuous variables are measured using t-tests and differences across factor variables are estimated using Pearson’s Chi-squared tests. While these statistical tests only reveal the nature of the association between the variables, they also serve to inform any subsequent modeling of the relationship between models of care and identified outcomes.

### Defining an episode of care

There is no universally agreed upon definition of an episode of care for these infection diagnoses. As noted above, in empirical work, researchers have defined episodes of different lengths. In Sweden, a national collaboration among general practitioners and the national public health agency has defined an episode of care for infections to be nominally 10 days, based on the concept of “infection episode” (Primärvårdskvalitet 2021). An episode starts on the day when an infection diagnosis, as defined by a set of ICD-10 infection codes, is identified and registered by a physician or a nurse at a specified clinic. This consultation is referred to as the index consultation. The index consultation needs to be preceded by a period of 10 days during which no diagnosis from the same category is registered. If an additional diagnosis of the same type is registered within 10 days of the index consultation, the episode continues. The episode ends after a period of 10 days during which no additional infection diagnosis of the same type has been registered. Consequently, an episode of care for infections in the current context is at least 10 days but may be longer in some cases. The identification of episodes requires access to patient identification data, contact date information, and diagnostic code by consultation. The data on the current sample enable the identification of episodes of care based on these criteria. In particular, the data enable the estimation of the number of episodes by patient, the length of episodes, and the types of consultations within an episode of care.

To assess the robustness of the main findings a number of additional analyses are performed. First, the length of the episode of care is extended correspondingly to 15 and 30 days, respectively. Second, relevant estimates are repeated for the specific infection types. Third, estimates are repeated also for other types of diagnoses. And finally, the sample is adjusted to control for any censoring effect.

## Results

The presentation of the findings of the analysis is organized along the specific objectives identified above. The first sub-section presents the results of estimating the number of consultations per episode of care by model of care (i.e., the intensity of care). The second sub-section reports the event of a switch in care model, both across the full sampling period and by a 10-day episode of care. The following sub-sections present the estimates of the content of care, including the share of consultations handled by a physician, the share of consultations involving a laboratory test, and the share of consultations resulting in an antibiotic prescription.

### Consultation intensity per episode of care

For the current sample of patients, the majority of cases would be expected to be managed within a single consultation. Table 2 shows the share of consultations that only involve a single visit or contact across the three models of care.

**Table 2.** Single-consultation episodes of care by type of consultation; frequency and percent

|  | Visit | Type of consultation |  | Total |
| --- | --- | --- | --- | --- |
|  |  | Digital | Telephone |  |
| Single-consultation episodes of care |  |  |  |  |
| No | 347,609 | 13,194 | 29,043 | 389,846 |
|  | 24.59% | 9.00% | 35.07% | 23.73% |
| Yes | 1,066,166 | 133,358 | 53,762 | 1,253,286 |
|  | 75.41% | 91.00% | 64.93% | 76.27% |

Source: Digital primary care study, DPC; consultation level data for sampling period. Telephone includes letter contacts.

Overall, around three-quarters of all 10-day episodes of care involve a single consultation, i.e., the consultation was preceded by a 10-day period during which no similar diagnosis was registered and followed by a similar period in which no consultation for the same group of diagnoses took place. Across the models of care, around 91 percent of all digital contacts only involved a single contact compared with 75 percent of the in-person visits and around 65 percent of the telephone contacts.

Restricting the sample to physician-led consultations that involved more than one consultation of the same type per 10-day episode of care, table 3 shows the mean number of consultations per such episodes across the three types of consultations.^9^

**Table 3.** Number of physician-led consultations per 10-day episodes of care by same type of consultation

|  | Visit | Type of consultation |  | Total |
| --- | --- | --- | --- | --- |
|  |  | Digital | Telephone |  |
| Mean, (SD) | 2.89 | 2.25 | 2.73 | 2.85 |
|  | (4.56) | (0.72) | (1.28) | (4.31) |

Source: Digital primary care study, DPC; consultation level data for sampling period; regional sample.
Sample includes only 10-day episodes of physician-led care involving multiple consultations of the same type. Telephone includes letter contacts.

For these types of episodes of care, the mean number of digital contacts was 2.25 compared with 2.89 in-person visits (with a standard deviation of 4.56) and 2.73 telephone contacts with a total average of around 2.85 consultations per 10-day episode of physician-led care of a similar type.

### Consultation switches across study period and within episodes of care

As noted, a particular objective of the analysis is to understand the possible association between the utilization of the alternative models of care. In particular, the possibility that the utilization of digital care affects the use of in-person care (or vice versa) would be particularly relevant to assess. While the above analysis showed that a number of patients made multiple in-person visits during the study period, the data also show that subsequent consultations sometimes involved a switch from one type of care to another. For patients that made more than one consultation during the full study period, around 85 percent of these involved no change in the type of consultation. Conversely, around 15 percent of patients used more than one type of care during the study period. Restricting the sample to 10-day episodes of care involving more than one consultation the number of patients switching from one type of care to another is relatively low, table 4.

**Table 4.** . Switch of consultation type; overall and within 10-day episodes of care; frequency and percent

|  |  |  |
| --- | --- | --- |
| Switch of consultation type, overall |  |  |
| No | 1,393,141 | 84.79% |
| Yes | 249,991 | 15.21% |
| Visit-to-Digital |  |  |
| No | 1,642,242 | 99.95% |
| Yes | 890 | 0.05% |
| Visit-to-Telephone |  |  |
| No | 1,625,206 | 98.91% |
| Yes | 17,926 | 1.09% |
| Digital-to-Visit |  |  |
| No | 1,641,208 | 99.88% |
| Yes | 1,924 | 0.12% |
| Digital-to-Telephone |  |  |
| No | 1,643,086 | 100.00% |
| Yes | 46 | 0.00% |
| Telephone-to-Visit |  |  |
| No | 1,636,802 | 99.61% |
| Yes | 6,330 | 0.39% |
| Telephone-to-Digital |  |  |
| No | 1,643,097 | 100.00% |
| Yes | 35 | 0.00% |

Source: Digital primary care study, DPC; consultation level data for sampling period.

With respect to a change from digital to in-person care or the other way around, the findings show that twice as many switched from digital to in-person compared with the opposite. However, the absolute numbers are small resulting in very small relative shares of the total number of consultations for either type of switch.

### Content of consultations

The third dimension of primary care effectiveness involves the content of the care. Specifically, three content measures of physician led care, involvement of laboratory testing, and prescription of antibiotic are estimated. Table 5 shows that digital contacts were delivered all but exclusively by a physician compared with the other two models of care. Somewhat over half of patients who made an in-person visit were seen by a physician while around three-quarters who made a telephone contact were treated by a physician.

**Table 5.**
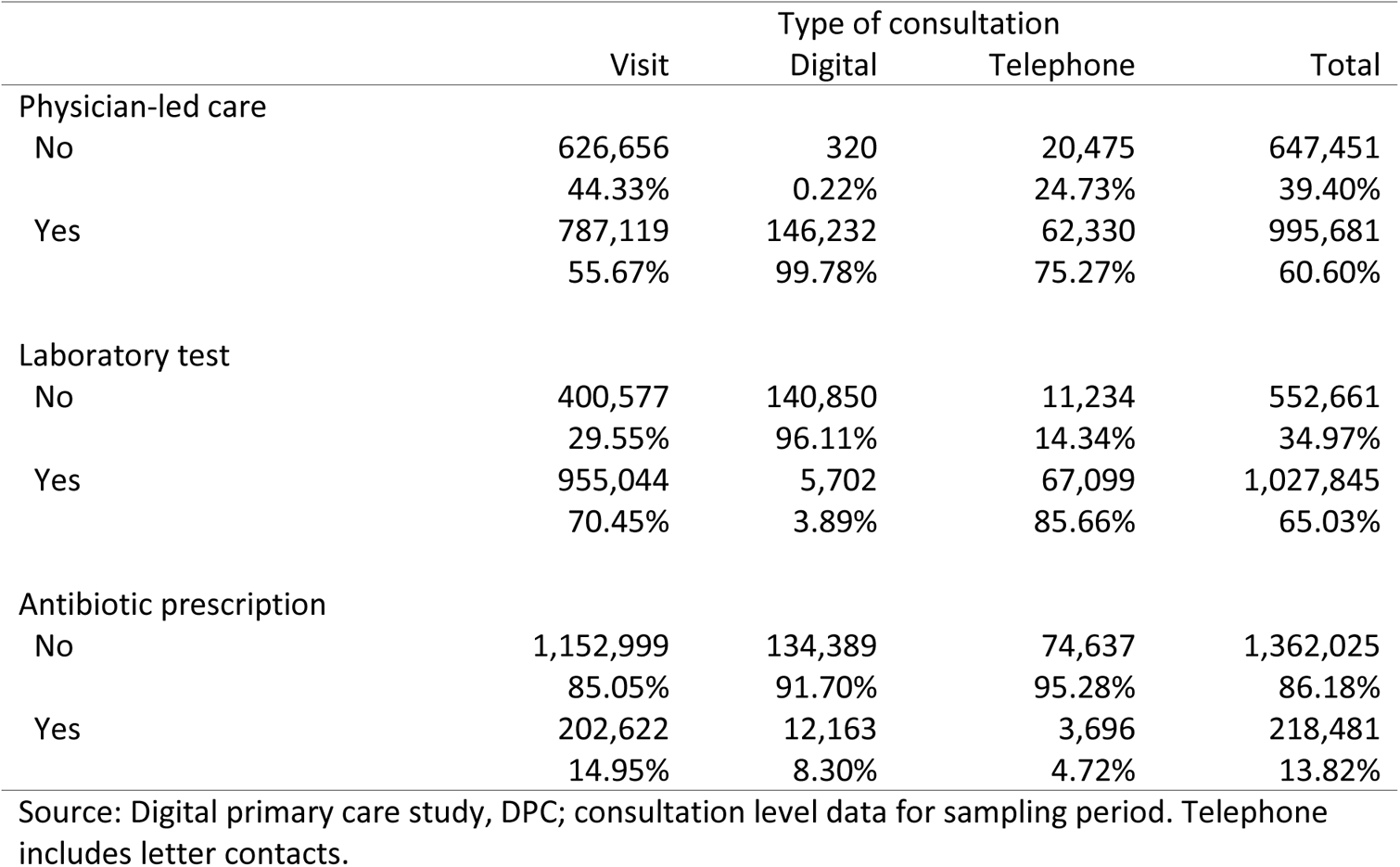
Physician-led care, laboratory test, and antibiotic prescription; frequency and percent

On the other hand, a considerably smaller share of consultations in digital care involves a laboratory test compared with in-person visits and telephone contacts. Similarly, antibiotic prescriptions were around half as common in digital care compared with in-person visits and twice as common compared with telephone contacts.

### Sensitivity analysis

The main results presented above suggest some differences between the alternative models of care as to their effectiveness in terms of the number of consultations per episode of care and the content of care. However, these results may be sensitive to variations in the identified underlying measures. To assess the extent to which the findings are robust to variations in the choice of measures or sample, this sub-section presents the results of some sensitivity analyses using variations in the length of care episodes, by specific infection type, and by the underlying sample. The results are presented in tables A.2a to A.6 in the Annex.

The results of the sensitivity analyses indicate that the main results are for the most part robust to these alternative specifications. First, while the extension of the episode of care period to 15 and 30 days, respectively, reduces the shares of single-consultation episodes it does so relatively equally across the three models of care (tables A.2.a and A.2.b). Similarly, the mean number of consultations of the same type increases when the episode of care periods are extended, but do so uniformly across the models of care (tables A.3.a and A.3.b). Extending the episodes of care to 15 and 30 days does not alter the main finding of digital care being the least intensive form of care in the current sample.

Second, looking at an episode of care over a 15-day period reduces the difference between a change in the model of care from digital to in-person visit and vice versa with slightly more changes moving from digital to in-person care (table A.4.a; the equivalent analysis for a 30-day episode is not shown for reasons of space). Third, the main results do not appear to be sensitive to the particular type of infection diagnosis as most values are distributed evenly across the infection types and models of care in terms of single-consultation episodes (table A.5, the same results are found when looking at multiple-consultation episodes, not shown). Fourth, the disaggregation of infection type does not change the overall finding with respect to switches of one type of care to another within a 10-day episode of care (table A.6). Nor does one type of infection appear to lead to a switch of consultation type to a larger extent than another type. Looking at the intensity of care and switches of care model within a 10-day episode of care using the full sample of patients diagnosed also with other conditions than an infection does not change the overall results of the main analysis (not shown). Finally, restricting the sample period to control for any censoring effect altered the results only marginally (not shown) as did removing observations with very large number of consultations (above 200 and above 100; not shown).

## Discussion

The introduction and expansion of new models of health care into public (or private) health systems can only be motivated by convincing evidence that they are effective, efficient, safe, and equitable. If new models fail to show sufficient effectiveness, it is difficult to argue for their inclusion and public subsidies. However, analyzing the effects of a particular model of care on policy relevant outcomes is difficult due to the complexities involved in the demand and supply of medical care that drive the data generating process. Based on a simple framework that outlines the choices individuals face when experiencing symptoms that may need some form of care the study finds both differences and similarities with respect to the effects of digital primary care compared with in-person care and telephone contacts.

### Intensity of care

In the current sample of patients diagnosed with an infection there is no indication that digital care requires more intense use of services compared with in-person visits or telephone contacts with clinics. A larger share of the digital care cases was managed in a single consultation compared with the former models. Likewise, for the multiple-consultation 10-day episodes of care, the mean number of consultations in the digital model was lower compared with the in-person model (and slightly lower compared with telephone contacts). However, further analysis shows that there are important differences between the models. In particular, the office-based model estimates are partly driven by a relatively small number of very high users of in-person care (see figure A.3). These are almost exclusively nurse-led cases, of which there are very few of in the digital model of care. This in turn indicates an important difference in the case-mix between in-person care and digital (and telephone) care where the former manages more severely ill patients that may require more intense levels of care over extended periods of time.

### Changes in model of care

One source of difference in effectiveness between the alternative models of care is that an episode of care is initiated in one model and continued in another as symptoms remain. In the current sample there is evidence of patients switching between models, both generally across the entire study period and within defined episodes of care (as illustrated in Figure 2). While the majority of patients have used only one type of care during the study period, some patients have switched type of consultation during the care episode. However, these changes of care model during an episode were relatively few, both in absolute numbers and as a share of the total number of consultations. In particular, less than one percent of consultations involved an episodic switch from in-person visits to digital care or vice versa. Looking at switches outside of the particular care episode these shares were even smaller (0.05 for a switch from in-person visits to digital contacts and 0.12 for the opposite switch). As noted above, any such effect of digital care on overall utilization of services would appear to capture a different data generating process more related to the preferences of the individuals as opposed to the effectiveness of the care provided. In the current sample, there would thus appear to be some support for the possibility that digital care leads to an increase in in-person visits, although the net effect is very small even in a large sample such as this one.

### Content of care

With respect to the content of care of the various models, the analyses of these measures indicate diverse, but relevant findings. First, digital care is exclusively provided by physicians in the current sample. As physician-led care is more costly compared with, e.g., nurse-led care, this would imply that digital care is less effective compared with in-person care and telephone contacts, all else equal.^10^ On the other hand, as digital care appears to manage cases by means of a single-consultation to a larger extent than in-person visits, the net effect may be smaller than the first measure would suggest. However, the previous findings suggested a difference in the case-mix between the models indicating that all else is not equal.

Second, the largest differences between the models were found with respect to the use of laboratory testing in the care process. Here, the analysis showed that digital care makes use of laboratory tests in less than four percent of cases compared with 70 percent and 86 percent in in-person care and telephone contacts, respectively. The significantly lower use of laboratory testing in digital care could be due to the very nature of the care model with less ability to conduct such tests. However, telephone contacts do involve a relatively large share of laboratory testing despite it also being a remote type of care. This is most likely a result of the fact that telephone contacts are made by patients registered with the particular health care clinic generating a more integrated care process as indicated by the number of patients switching from in-person visits to telephone contacts within the episode of care (see Table 4). It may also reflect a difference in case-mix between the models of care.

The third finding with regard to the content of the care showed that digital care results in an antibiotic prescription around half as frequently as in-person care (and twice as frequently as telephone contacts). Again, all else equal, digital care appears not to be less effective compared with in-person care for these types of diagnoses and, furthermore, does not appear to lead to an over-use of antibiotics in primary care.^11^

### Additional findings

In addition to the specific findings described above, an additional finding is that the utilization of primary care services in the current sample is highly skewed (18.1 across the full sample). Around 50 percent of the individuals in the sample make only one consultation over the entire study period. An additional 22 percent make two consultations during the period and another 11 percent make three consultations. Of those who make more than one consultation, around 15 percent make use of more than one type of model of care over the entire period (see Table 4). As digital care and new forms of primary care services more generally (such as retail care and other types of clinics (Pollack, Gidengil et al. 2010, Hoff and Prout 2019)) expand the group of patients using more than one type of care will most likely grow and it will be increasingly more difficult to speak of ‘users’ of a particular type of service. Understanding these changes in utilization behaviors will be important for effective health care policy designs and regulations.

Moreover, for most cases in either model of care for these conditions, patients are referred to the self-care model of care without a laboratory investigation or a prescription for antibiotics (Figure 2). In practice, most patients are advised to rest and possibly make use of non-prescription drugs and to return in case symptoms remain beyond a certain time period. The data also suggest that in the majority of cases, this is the appropriate management of these conditions as relatively few cases require follow-up consultations.

The finding that most cases are referred to the self-care model does not necessarily suggest that the majority of consultations for this group of patients were medically unnecessary. However, it may suggest that primary care systems are facing two related and partially conflicting patient management problems. First, given the fact that most patients in either the in-person or the digital model of care are screened in a triage procedure before being seen by a physician, primary care systems are facing a matching problem where too many patients end up being seen by a physician only to be referred to self-care without further medical intervention. And second, such systems also face a risk management problem involving the identification of patients that do require further investigation and possibly need antibiotic treatment and those who do not. It is possible that digital technologies for health care can contribute to handling both of these problems by offering the ability to match patients to a more cost-effective type of care and by collecting relevant information to support clinical decisions. In turn, that may require the ability to combine digital care with the possibility to perform laboratory tests to strengthen this identification process for this group of patients.

Furthermore, from a health systems perspective, this has implications for how these kinds of conditions should be managed. To the extent that digital care is less costly in terms of direct and indirect resource use (including time) from the perspective of both providers and patients, these conditions should be managed by means of digital care. However, being able to identify cases that may need to be seen by a physician, is difficult *a priori* (Entezarjou, Bonamy et al. 2020). To be able to perform such identifications, health systems need to be fully integrated, either organizationally or virtually through common medical records and communication technologies for sharing patient data. Doing so in a safe and ethical manner will be critical for the successful introduction of digitally based care (Coyle, Diepeveen et al. 2020).

### Relationship with the current evidence

The findings of the study contrast with some of the results in other studies. For example, two recent studies that have used similar data from the same context as the current study report that digital care lead to an overall increase in primary care consultations (Ellegård and Kjellsson 2019, Ellegård, Kjellsson et al. 2021). However, these two studies have used different outcome measures of the effects of digital care, which may explain these differences. Specifically, the current study has focused on the effects of digital care related to an episode of care while the other two studies have looked at the effect of digital care on the universe of consultations. As discussed above, the latter approach is challenging if the sample does not include the true universe and also can control for any external change to the data collection process. While the use of a particular model of care at some point may affect the choice at some later point outside of the episode of care, it is most likely not the effect of the particular type of care as such, but rather the perceptions of the individuals that explain the outcome. While the perceptions of individuals are also reflected in the choice of care over an episode of care, it is more likely that the episodic approach more directly measures the effectiveness of digital care.

### Contributions to the existing evidence base

The findings of this study add to the existing body of evidence on the impacts of digital care. First, the analysis is based on an analytical framework that describes the data generating process underpinning the approach and the identification of the outcome measures. This facilitates the interpretation of the findings and reinforces the understanding of the analysis. Second, it makes use of a large dataset with detailed information on the individuals and on the characteristics of the consultations, as well as on relevant process outcomes of the care. The sample covers one-third of the country in terms of regions and include both large and small regions. And third, it is able to identify episodes of care that conform to the formal guidelines on the treatment of patients with these infection diagnoses. This ability enhances the policy relevance of the analysis.

### Implications

There are a number of normative implications of this study. Over the coming years, it is likely that digital models of care will continue to be developed and introduced into existing health care systems, also in other medical fields, such as physiotherapy, mental health care, and cardiovascular diseases. In this process, policy makers and health authorities need to find the balance between fostering innovation and value for money. Given cost-effectiveness and patient safety concerns, digital care models need to be seen as viable alternatives for various groups of patients to access effective care. As such, all models of care, regardless of ownership, need to be fully integrated into existing systems of care, including by means of technology neutral reimbursement schedules. Such reimbursement systems need to both foster innovation and to focus on patient outcomes rather than inputs and quantity of services, which may or may not be effective. Finally, the onset of the Covid-19 pandemic may have led to a shift in the application of digital care, not just in the level of care, but also in the scope of primary care (Ekman, Arvidsson et al. 2021). The impact of such an external shock may also lead to a change in the effectiveness of digital care on identified outcomes as more providers become familiar with the technology and the nature of providing care by such means.

### Limitations

Although the study has made use of a large sample with detailed information on relevant factors related to primary care, there are a number of limitations of the study. First, the data do not contain outcome measures of the primary care process. For example, there is no information on when patients become free from symptoms after the first consultation when the diagnosis was set. Such kinds of data would be valuable but are rarely readily available for large samples of patients. Second, there is no information on whether patients were correctly diagnosed in either type of care. Such information is also not readily available for large samples. And third, the study includes patients diagnosed with an infection of some type. While this group of patients constitutes a large share of all primary care consultations, there are many other types of diseases and conditions, including those mentioned above, where digital care also is used. The effects of digital care for those types of conditions may be different from those found in the current study.

## Conclusions

Primary care provided by means of some digital technology, such as video or chat, may be a viable alternative model of care for the majority of patients diagnosed with an infection. Compared with the traditional models of primary care, digital care does not appear to be associated with more intense levels of care or lead to additional care in subsequent consultations within a defined episode of care. As digital care appears to differ from traditional models of care with regard to the content of care, further analyses are needed to estimate the cost-effectiveness of digital care compared with existing models of care. Further investigation is also needed to provide evidence on the long-term impact of digital care on overall utilization of care and on the appropriateness of digital care for particular patient groups. These analyses will need to take into account the non-random nature of the implementation of digital care in most health systems.

## Annex: Additional figures and tables

**Figure A.1:**
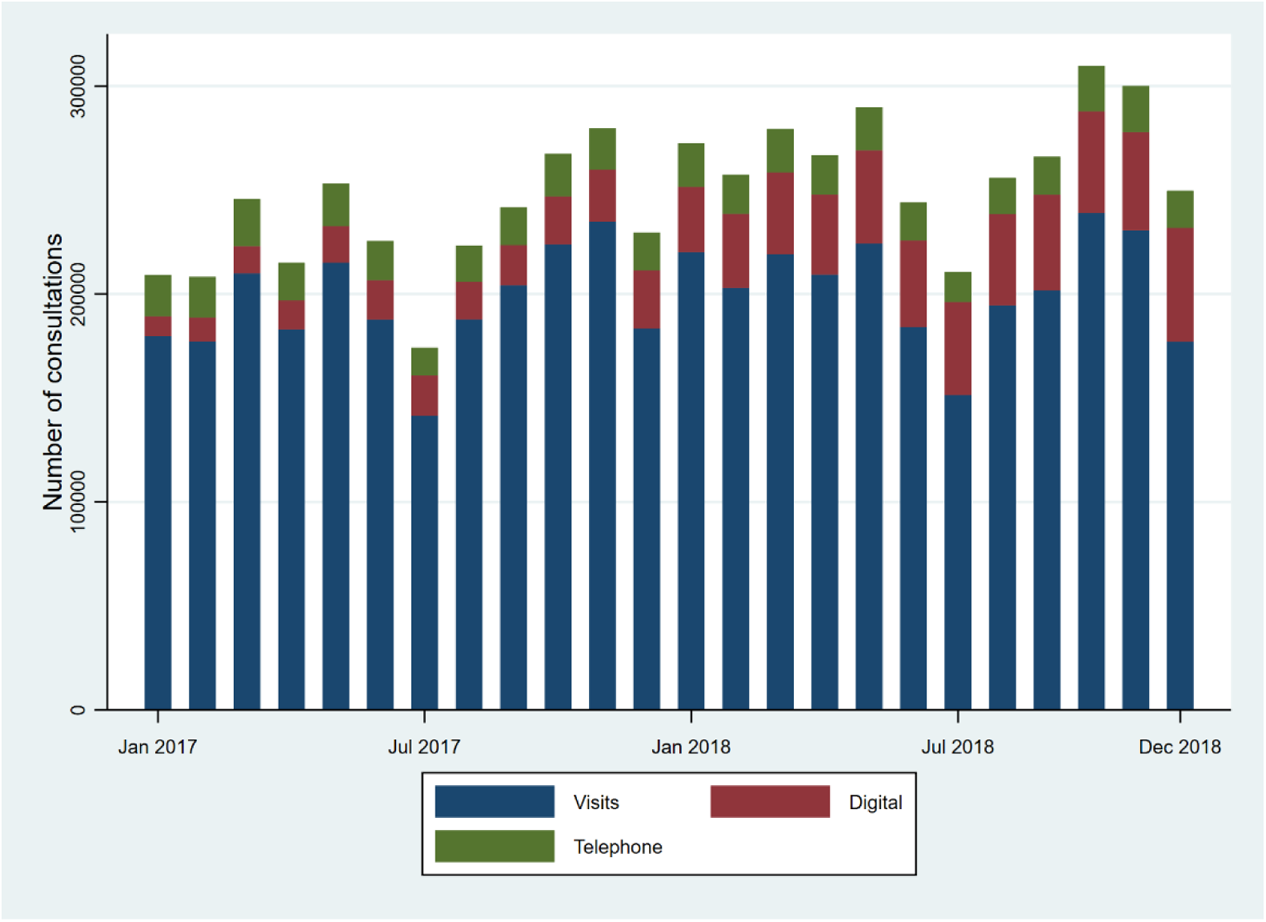
Consultations by month and type

**Figure A.2:**
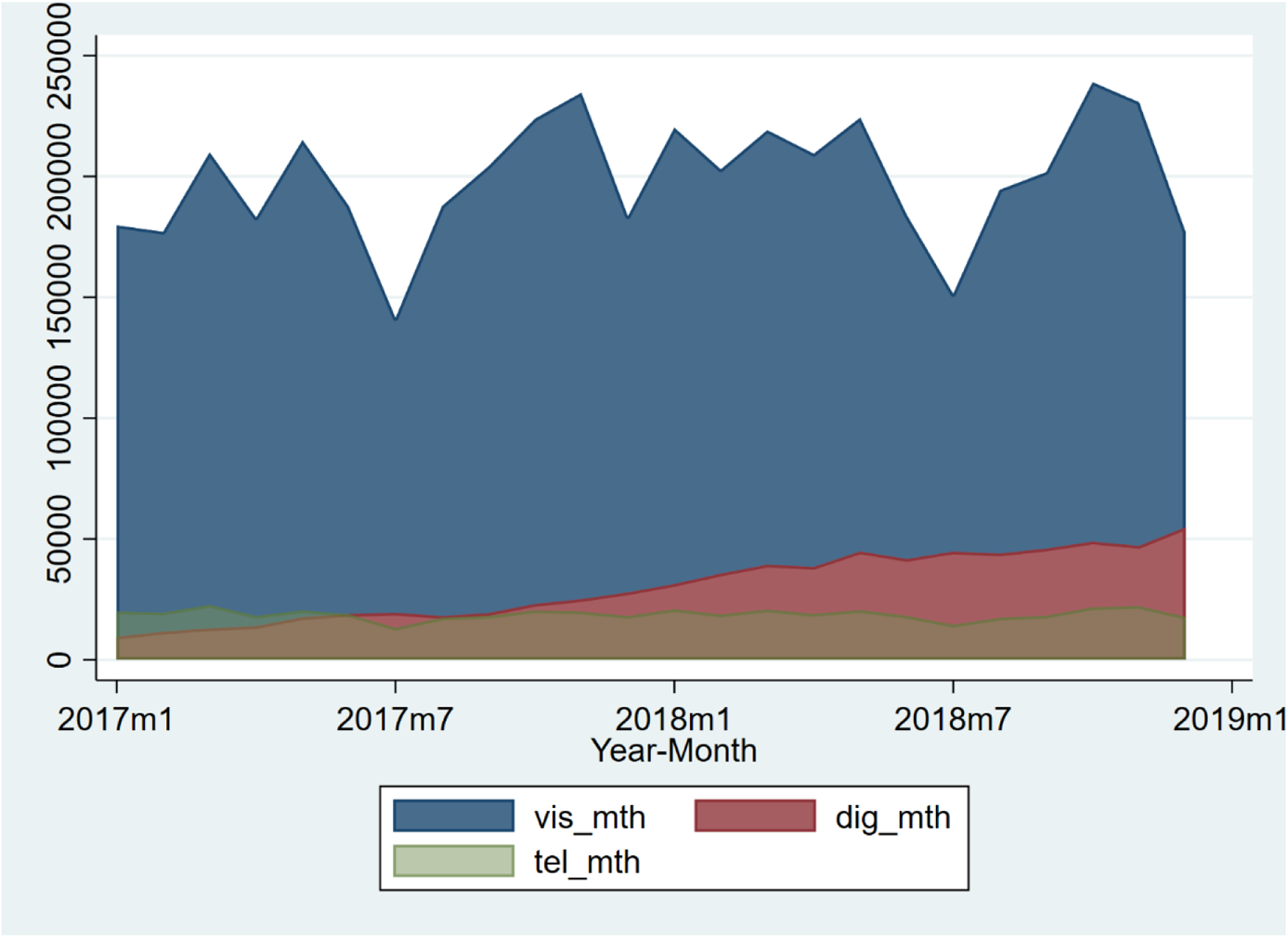
Consultation shares by type

**Figure A.3:**
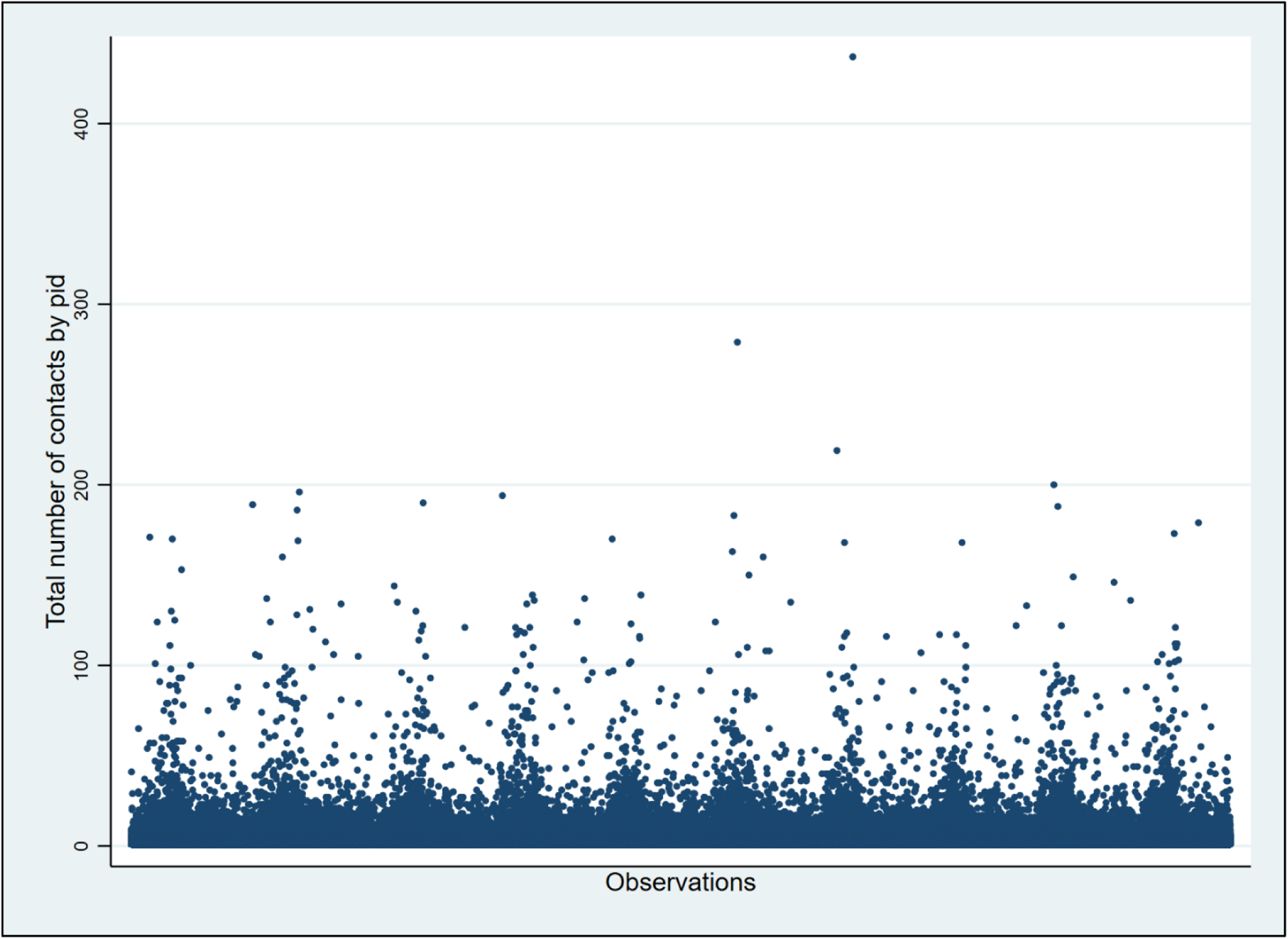
Number of consultations by individual

**Figure A.4.**
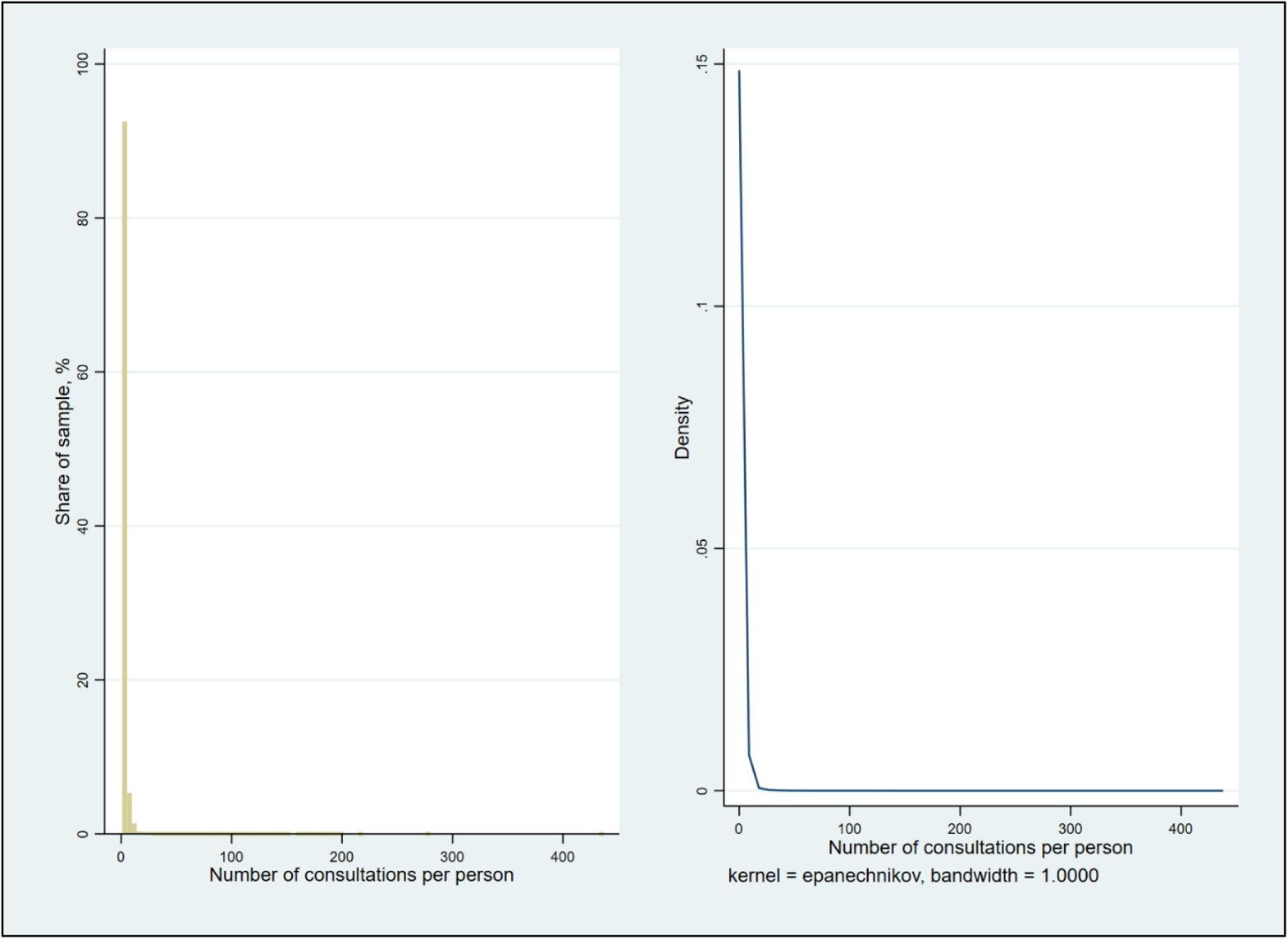
Distribution of consultations by individual (histogram, LHS; density plot, RHS)

**Table A.1.** Descriptive statistics of individual level sample by type of consultation and total

|  | Type of consultation |  |  | Total |
| --- | --- | --- | --- | --- |
|  | Visit | Digital | Telephone |  |
| Mean age, years | 51.47<br>(21.05) | 35.36<br>(12.97) | 58.80<br>(20.73) | 49.34<br>(20.92) |
| Mean annual gross income, SEK | 212,029<br>(246,467) | 293,436<br>(907,543) | 217,664<br>(258,855) | 223,954<br>(414,787) |
| Charlson comorbidity index, CCI | 0.13<br>(0.44) | 0.05<br>(0.29) | 0.19<br>(0.60) | 0.12<br>(0.43) |
| Sex |  |  |  |  |
| Female | 316,777<br>56.78% | 64,337<br>66.04% | 11,150<br>60.69% | 392,264<br>58.22% |
| Male | 241,151<br>43.22% | 33,085<br>33.96% | 7,221<br>39.31% | 281,457<br>41.78% |
| Education level |  |  |  |  |
| Elementary | 131,104<br>24.14% | 10,866<br>11.38% | 5,072<br>28.19% | 147,042<br>22.40% |
| High-school | 244,202<br>44.97% | 35,647<br>37.34% | 7,783<br>43.26% | 287,632<br>43.81% |
| University | 167,704<br>30.88% | 48,963<br>51.28% | 5,138<br>28.56% | 221,805<br>33.79% |
| Born in Sweden |  |  |  |  |
| No | 103,202<br>18.50% | 13,164<br>13.51% | 2,813<br>15.31% | 119,179<br>17.69% |
| Yes | 454,726<br>81.50% | 84,258<br>86.49% | 15,558<br>84.69% | 554,542<br>82.31% |
| Married |  |  |  |  |
| No | 314,176<br>56.31% | 66,535<br>68.30% | 9,828<br>53.50% | 390,539<br>57.97% |
| Yes | 243,752<br>43.69% | 30,887<br>31.70% | 8,543<br>46.50% | 283,182<br>42.03% |
| Employment status |  |  |  |  |
| Not employed | 256,093<br>45.90% | 17,239<br>17.70% | 10,069<br>54.81% | 283,401<br>42.07% |
| Employed | 301,835<br>54.10% | 80,183<br>82.30% | 8,302<br>45.19% | 390,320<br>57.93% |
| Chronic illness |  |  |  |  |
| No | 190,160<br>71.67% | 470<br>94.19% | 2,166<br>82.17% | 192,796<br>71.82% |
| Yes | 75,161 | 29 | 470 | 75,660 |
|  | 28.33% | 5.81% | 17.83% | 28.18% |
| Total | 557,928 | 97,422 | 18,371 | 673,721 |

Mean, frequencies, and shares. Standard deviation in parenthesis. 100 SEK = approx. \$11.6.
Source: Digital primary care study, DPC; individual level data for sampling period; regional sample.
Telephone includes letter contacts. Total number of individuals = 673,721.

**Table A.2.a.** Single-consultation episodes of care by type of consultation; 15-day

| Table A.2.2a: Single consultation episodes of care by type of consultation, 15-day |  |  |  |  |
| --- | --- | --- | --- | --- |
|  | Visit | Type of consultation |  | Total |
|  |  | Digital | Telephone |  |
| Single-consultation episodes of care; 15-day |  |  |  |  |
| No | 426,195 | 16,980 | 36,113 | 479,288 |
|  | 30.15% | 11.59% | 43.61% | 29.17% |
| Yes | 987,580 | 129,572 | 46,692 | 1,163,844 |
|  | 69.85% | 88.41% | 56.39% | 70.83% |

Source: Digital primary care study, DPC; consultation level data for sampling period. Telephone includes letter contacts.

**Table A.2.b.** Single-consultation episodes of care by type of consultation; 30-day

| Table A.2.1b: Single consultation episodes of care by type of consultation, 30-day |  |  |  |  |
| --- | --- | --- | --- | --- |
|  | Visit | Type of consultation |  | Total |
|  |  | Digital | Telephone |  |
| Single-consultation episodes of care; 30-day |  |  |  |  |
| No | 559,598 | 25,040 | 47,349 | 631,987 |
|  | 39.58% | 17.09% | 57.18% | 38.46% |
| Yes | 854,177 | 121,512 | 35,456 | 1,011,145 |
|  | 60.42% | 82.91% | 42.82% | 61.54% |

Source: Digital primary care study, DPC; consultation level data for sampling period. Telephone includes letter contacts.

**Table A.3.a.** Number of consultations per 15-day episodes of care by a similar type of consultation

|  | Visit | Type of consultation |  | Total |
| --- | --- | --- | --- | --- |
|  |  | Digital | Telephone |  |
| Mean, (SD) | 9.15 | 2.64 | 3.24 | 8.84 |
|  | (20.31) | (1.43) | (2.01) | (19.84) |

Source: Digital primary care study, DPC; consultation level data for sampling period; regional sample.
Sample includes only 15-day episodes of care involving multiple consultations of the same type.
Telephone includes letter contacts.

**Table A.3.b.**
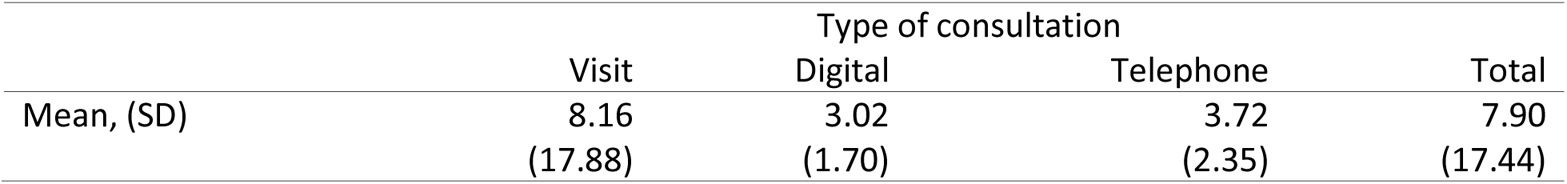
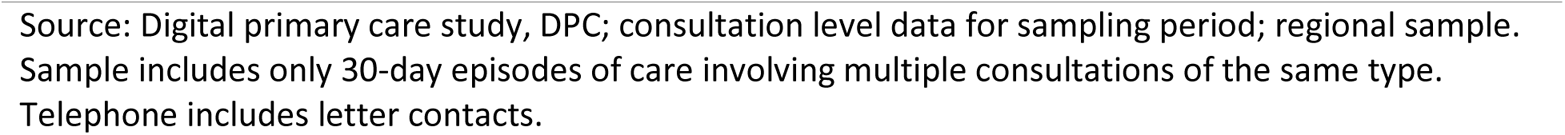
Number of consultations per 30-day episodes of care by a similar type of consultation

|  | Visit | Type of consultation |  | Total |
| --- | --- | --- | --- | --- |
|  |  | Digital | Telephone |  |
| Mean, (SD) | 8.16 | 3.02 | 3.72 | 7.90 |
|  | (17.88) | (1.70) | (2.35) | (17.44) |

Source: Digital primary care study, DPC; consultation level data for sampling period; regional sample. Sample includes only 30-day episodes of care involving multiple consultations of the same type. Telephone includes letter contacts.

**Table A.4.** Switch of consultation types within 15-day episodes of care

|  |  |  |
| --- | --- | --- |
| Visit-to-Digital; 15-day |  |  |
| No | 1,641,799 | 99.92% |
| Yes | 1,333 | 0.08% |
| Visit-to-Telephone; 15-day |  |  |
| No | 1,621,021 | 98.65% |
| Yes | 22,111 | 1.35% |
| Digital-to-Visit; 15-day |  |  |
| No | 1,640,836 | 99.86% |
| Yes | 2,296 | 0.14% |
| Digital-to-Telephone; 15-day |  |  |
| No | 1,643,070 | 100.00% |
| Yes | 62 | 0.00% |
| Telephone-to-Visit; 15-day |  |  |
| No | 1,634,689 | 99.49% |
| Yes | 8,443 | 0.51% |
| Telephone-to-Digital; 15-day |  |  |
| No | 1,643,090 | 100.00% |
| Yes | 42 | 0.00% |

Source: Digital primary care study, DPC; consultation level data for sampling period.

**Table A.5.** Single-consultation episodes of care by type of consultation and infection type

|  | Type of consultation |  |  |  |  |  |  |  |  |
| --- | --- | --- | --- | --- | --- | --- | --- | --- | --- |
|  | Visit |  |  | Digital |  |  | Telephone |  |  |
|  | Infection type |  |  | Infection type |  |  | Infection type |  |  |
|  | 1. RTI | 2. UTI | 3. SSI | 1. RTI | 2. UTI | 3. SSI | 1. RTI | 2. UTI | 3. SSI |
| Single-consultation episodes of care |  |  |  |  |  |  |  |  |  |
| No | 184,925 | 30,026 | 132,658 | 7,934 | 2,101 | 3,159 | 20,501 | 5,483 | 3,059 |
|  | 20.45% | 18.65% | 38.05% | 10.36% | 6.54% | 8.35% | 37.08% | 29.94% | 33.22% |
| Yes | 719,263 | 130,945 | 215,958 | 68,661 | 30,014 | 34,683 | 34,784 | 12,830 | 6,148 |
|  | 79.55% | 81.35% | 61.95% | 89.64% | 93.46% | 91.65% | 62.92% | 70.06% | 66.78% |

Source: Digital primary care study, DPC; consultation level data for sampling period. Telephone includes letter contacts.
RTI: Respiratory tract infection; UTI: Urinary tract infection; SSI: Skin- and soft-tissue infection.

**Table A.6.** Switch of consultation types by infection type; overall and within 10-day episodes of care

|  | Infection type |  |  |
| --- | --- | --- | --- |
|  | 1. RTI | 2. UTI | 3. SSI |
| Switch of consultation type, general |  |  |  |
| No | 872,120<br>84.18% | 172,366<br>81.54% | 348,655<br>88.12% |
| Yes | 163,948<br>15.82% | 39,033<br>18.46% | 47,010<br>11.88% |
| Visit-to-Digital |  |  |  |
| No | 1,035,394<br>99.93% | 211,323<br>99.96% | 395,525<br>99.96% |
| Yes | 674<br>0.07% | 76<br>0.04% | 140<br>0.04% |
| Visit-to-Telephone |  |  |  |
| No | 1,022,873<br>98.73% | 208,760<br>98.75% | 393,573<br>99.47% |
| Yes | 13,195<br>1.27% | 2,639<br>1.25% | 2,092<br>0.53% |
| Digital-to-Visit |  |  |  |
| No | 1,034,751<br>99.87% | 211,167<br>99.89% | 395,290<br>99.91% |
| Yes | 1,317<br>0.13% | 232<br>0.11% | 375<br>0.09% |
| Digital-to-Telephone |  |  |  |
| No | 1,036,042<br>100.00% | 211,390<br>100.00% | 395,654<br>100.00% |
| Yes | 26<br>0.00% | 9<br>0.00% | 11<br>0.00% |
| Telephone-to-Visit |  |  |  |
| No | 1,031,595<br>99.57% | 210,344<br>99.50% | 394,863<br>99.80% |
| Yes | 4,473<br>0.43% | 1,055<br>0.50% | 802<br>0.20% |
| Telephone-to-Digital |  |  |  |
| No | 1,036,045<br>100.00% | 211,391<br>100.00% | 395,661<br>100.00% |
| Yes | 23<br>0.00% | 8<br>0.00% | 4<br>0.00% |

Source: Digital primary care study, DPC; consultation level data for sampling period.
RTI: Respiratory tract infection; UTI: Urinary tract infection; SSI: Skin- and soft-tissue infection.

## Data Availability

All data produced in the present work are contained in the manuscript.

## Footnotes

1 There may also be a novelty factor with respect to digital care, which also may affect individuals’ choice with respect to care models.

2 Quality-Adjusted Life-Years (QALYs) and similar composite health outcome measures of health care treatments are rarely available in the general primary health care setting. Even more direct measures of the effectiveness of the treatments of common conditions in primary care, such as reductions in fever or levels of symptoms, are usually not available in register data.

3 See, for example, Chapter 2 in Phelps (2018) for the formal representation of the individual’s maximization problem in health care.

4 I.e., once care has been initiated, the individual has little or no ability to affect the scope or content of the provider-led care by means of her behavior.

5 As noted, register data on consultations rarely provide sufficient information to assess the medically appropriateness of the particular consultation. Patient records data may provide such information.

6 It may also be noted that neither approach is able to distinguish between care that can be defined as medically “necessary” and care that can be defined as “unnecessary” for the reasons discussed above. Importantly, neither model is able to estimate the absolute effects of care due to the lack of a control (un-treated) group; in RCT-terms, they are A/B-studies with two (or more) treatment arms.

7 In the total sample, individuals from all 290 municipalities of Sweden are included. However, to avoid any risk of data leakage, observations from outside the seven sampling regions are dropped in the analyses of episodes of care as patients who made a digital contact could have made an in-person visit in their home region outside of the sampling regions.

8 These context indicators are not used in the current analysis.

9 As described above, an episode of care may involve more than 10 days if a new consultation for a similar diagnosis is registered within 10 days.

10 In particular, it is an indication of a difference in the cost-effectiveness of the care.

11 It is not possible from this analysis to conclude whether either of the model of care is effective (or cost-effective) in an absolute sense.

## Notes

### Competing Interest Statement

The authors have declared no competing interest.

### Funding Statement

This study was funded by the Swedish Research Council for Health, Working Life, and Welfare (Forte; grant number 2018-00093).

### Author Declarations

The Swedish Ethical Review Authority gave ethical approval for this work (2019-01500; 2019-03-20).

